# Evaluation of the Causal Relationship Between Smoking and Schizophrenia in Asia

**DOI:** 10.1101/2022.03.07.22272059

**Authors:** Mei-Hsin Su, Rou-Yi Lai, Yen-Feng Lin, Chia-Yen Chen, Yen-Chen A. Feng, Po-Chang Hsiao, Shi-Heng Wang

**Author notes:** Corresponding author. College of Public Health, China Medical University, Taichung, Taiwan., *Email address* (S.H. Wang).

## Abstract

Cigarette smoking has been suggested to be associated with the risk of schizophrenia (SCZ) in observational studies. A significant causal effect of smoking on SCZ has been reported in the European population using the Mendelian randomization (MR) approach; however, no evidence of causality was found in participants from East Asia (EAS). Using the Taiwan Biobank (TWBB, sample size up to 79,989), we conducted genome-wide association studies (GWAS) to identify susceptibility loci for smoking behavior, which included the initiation of smoking and the onset age. To maximize the power of genetic discovery in the EAS population, we meta-analyzed GWAS from the TWBB and Biobank Japan (BBJ, sample size up to 165,436) for smoking traits. The GWAS for SCZ was taken from the Asia Psychiatric Genomics Consortium, which included 22,778 cases and 35,362 controls. We performed a two-sample MR to estimate the causality of smoking behavior on SCZ in the EAS population. In TWBB, we identified one novel locus that met genome-wide significance for onset age. In a meta-analysis of TWBB and BBJ, we identified two novel loci for smoking initiation. In MR, a marginal significance was found for the causality of smoking initiation on SCZ (odds ratio (OR) = 4.00, 95% confidence interval (CI) = 0.89–18.01, *P* = 0.071). Later onset age for smoking was causally associated with a lower risk of SCZ (OR for a per-year increase in onset = 0.96, 95% CI = 0.91–1.01) with a marginal significance (*P* = 0.098).

## 1. Introduction

Schizophrenia (SCZ) is often comorbid with smoking behaviors. Both case-control and cohort studies indicated that smokers were at a significantly higher risk of SCZ than non-smokers (Weiser et al., 2004), especially heavy-smokers (Hunter et al., 2020). Meta-analyses, including cohort studies, also indicated that smokers have a two-fold increased relative risk of SCZ than non-smokers (Gurillo et al., 2015; Hunter et al., 2020). In addition, the comorbidity of smoking and SCZ is associated with poorer prognosis (Whiteford et al., 2013) and aggravated mental symptoms, such as flat affect, delusions, and hallucinations (Kelly & McCreadie, 1999).

Some possible explanations have been proposed for the association between smoking behaviors and SCZ. One argument is that substance use may lead to SCZ. According to reports, 90% of patients with SCZ started to smoke before the onset of their illness (Kelly & McCreadie, 1999), and adolescents with smoking behaviors were more likely to develop SCZ than non-smoking adolescents (de Leon, 1996; Weiser et al., 2004). These studies used temporal sequence approach to infer the possibility that substance use may lead to SCZ. Another possible reason is shared genetic architecture. In our previous study, we calculated the polygenetic risk scores of SCZ based on genome-wide association studies (GWAS), and found that the score was positively associated with lifetime tobacco smoking (Wang et al., 2019). On the other hand, the polygenic risk scores of cotinine concentrations also significantly predict schizophrenia diagnosis (Chen et al., 2016). In addition, a genetic correlation was found between SCZ and regular smoking (Harper et al., 2021). Self-medication is another hypothesis for the association between smoking and SCZ. Patients with SCZ may try to diminish their symptoms and the side effects of antipsychotic drugs through nicotine consumption, which can increase hepatic clearance and restore the dopamine blockade (Winterer, 2010). Animal models also provided evidence that chronic nicotine administration reversed hypofrontality by nicotinic acetylcholine receptor (nAChR) modulation and further attenuated the symptoms of SCZ (Koukouli et al., 2017; Noda et al., 2020).

The association between smoking and SCZ has been established, but the causal relationship between them remains conflicting. Investigating the causal relationship between smoking and SCZ may help in blocking the path to SCZ and subsequently reduce the disease burden. The Mendelian randomization (MR) is a novel study design for investigating causality using a genetic approach. Reports suggest that smoking plays a causal role in SCZ (Barkhuizen et al., 2021; Gage et al., 2017; Wootton et al., 2020; Yuan et al., 2020). Further, the causal effect of SCZ on smoking initiation behaviors was also detected (Wootton et al., 2020). However, most studies were conducted in Caucasian populations with a sufficient sample size for GWASs for smoking and SCZ. Only one MR analysis from the Asian population did not support such causality (Chen et al., 2021). The negative finding may have resulted from insufficient power for the MR analysis with a limited sample size for GWAS. Therefore, estimating the causal relationship with a larger sample size in the Asian population can help provide precise results.

Since smoking behaviors are associated with various physical (Moudi et al., 2018; Teramoto et al., 2021) and mental illnesses (Hunter et al., 2020; Wang et al., 2019), investigating the causality between them helps block the pathway which may lead to SCZ. Therefore, this study aimed to conduct a well-powered GWAS for smoking initiation and onset age among East Asian populations with larger sample size which came from meta-analyses of the Taiwan Biobank (TWBB) and Biobank Japan (BBJ). We then examined the causal relationship between these two smoking behaviors and SCZ in the Asian population.

## 2. Method

### 2.1. GWAS for smoking

This study used individual genotyping and phenotyping data from the TWBB, the largest government-supported biobank in Taiwan since 2012. The TWBB recruits community-based samples aged 30–70 years who are cancer-free at recruitment. The internal review board approved the recruitment and data collection procedures of TWBB. Each participant signed an approved informed consent form, provided blood samples, and underwent physical examinations and face-to-face interviews. This study was approved by the Central Regional Research Ethics Committee of China Medical University, Taichung, Taiwan (CRREC-108-30).

Genotyping of 95,238 TWBB participants was performed using customized TWBB chips and processed on the Axiom Genome-Wide Array Plate System (Affymetrix, Santa Clara, CA, USA). Furthermore, 26,274 participants were genotyped on the TWBv1 chip, and 68,964 participants were genotyped on the TWBv2 chip. We conducted quality control and imputation of the two chips separately. Quality control included the exclusion criteria of variants with call rate < 95%, individuals with more than 5% missing variants, minor allele frequency (MAF) < 0.001, and deviation from Hardy-Weinberg equilibrium with *P* < 1 × 10^−6^. Imputation was performed based on the 973 TWBB panels from whole-genome sequencing in TWBB participants and 504 EAS panels in the 1000 Genomes project, variants with MAF ≥ 0.5%, and imputation INFO score ≥ 0.7. In addition, cryptic relatedness was removed, and we estimated identity by descent (IBD) sharing coefficients, PI-HAT = probability (IBD = 2) + 0.5 × probability (IBD = 1), between any two participants in KING and excluded one individual from a pair with PI-HAT ≥ 0.1875.

GWAS for two smoking behaviors included smoking initiation (yes/no) and onset age in a sample size of 79,989 individuals. We performed linear or logistic regression in PLINK for association tests with adjustment for age, age^2^, sex, age by sex interaction, age^2^ by sex interaction, and the top 20 principal components. In addition, we performed a genetic association test separately for the two chips and performed an inverse-variance-weighted fixed-effect meta-analysis in METAL. For each trait, the sample size and heritability estimates for each chip and genetic correlation estimates (Bulik-Sullivan et al., 2015a, 2015b) between the two chips are detailed in Supplementary Table 1.

**Table 1.** Genome-wide association analysis for smoking behavior in Asia population

| Lead SNP | chr | Position<br>(hg38) | Effect<br>allele | gene | TWBB |  |  | BBJ |  |  | Combined |  |  |
| --- | --- | --- | --- | --- | --- | --- | --- | --- | --- | --- | --- | --- | --- |
|  |  |  |  |  | Beta | SE | p | Beta | SE | p | Beta | SE | p |
| Smoking initiation |  |  |  |  |  |  |  |  |  |  |  |  |  |
| rs79574881 | 8 | 13420362 | A | *DLC1* | -0.06 | 0.046 | 0.2039 | -0.02 | 0.004 | 1.10E-07 | -0.02 | 0.004 | **2.84E-08** |
| rs2156008 | 18 | 55614013 | A | *TCF4* | 0.06 | 0.014 | 7.90E-06 | 0.01 | 0.002 | 5.30E-07 | 0.01 | 0.002 | **3.21E-08** |
| Onset |  |  |  |  |  |  |  |  |  |  |  |  |  |
| rs553874586 | 17 | 55405197 | A | *MMD* | -3.77 | 0.654 | **8.08E-09** | - | - | - | -3.77 | 0.654 | **8.07E-09** |

To maximize the power of genetic discovery in East Asia, we meta-analyzed GWAS from the TWBB and BBJ (sample size up to 165,436, for each smoking trait with MAF ≥ 0.5% and INFO score ≥ 0.7) (Matoba et al., 2019), including smoking initiation (N = 245,425) and onset age (N = 46,000). The genetic correlation between TWBB and BBJ was median to high for smoking initiation but not for onset age (Supplementary Table 2).

**Table 2.** The causal estimation of smoking behaviors on schizophrenia in Asia population

| Exposure | N<br>SNP | MR Method | N<br>outliers | OR (95% CI) | p-value |
| --- | --- | --- | --- | --- | --- |
| Smoking initiation | 8 | IVW |  | 4.00 (0.89-18.01) | 0.0714 |
|  |  | Weighted median |  | 3.44 (0.56-21.11) | 0.1826 |
|  |  | Weighted mode |  | 4.57 (0.30-69.27) | 0.3091 |
|  |  | MR-PRESSO | 0 | 4.00 (1.78-9.98) | 0.0122 |
|  |  | MR Egger |  | 4.14 (0.01- >99) | 0.6562 |
|  |  | (intercept, SE) |  | -0.0003 (0.0281) | 0.9906 |
| Onset | 5 | IVW |  | 0.96 (0.91-1.01) | 0.0982 |
|  |  | Weighted median |  | 0.97 (0.91-1.03) | 0.5071 |
|  |  | Weighted mode |  | 0.97 (0.90-1.05) | 0.3311 |
|  |  | MR-PRESSO | 0 | 0.96 (0.91-1.01) | 0.1736 |
|  |  | MR Egger |  | 0.93 (0.83-1.05) | 0.3496 |
|  |  | (intercept, SE) |  | 0.0170 (0.0367) | 0.6757 |

### 2.2. GWAS summary for SCZ

The GWAS summary of SCZ from the Asia Psychiatric Genomics Consortium (PGC), including 22,778 cases and 35,362 controls (Lam et al., 2019) from East Asia, and the diagnosis of SCZ was based on the Diagnostic Manual of Mental Disorders-IV (DSM-IV) system.

### 2.3. Mendelian randomization

#### Genetic instrument for causality of smoking on SCZ

We mapped the variants from exposure (smoking phenotypes) data to outcome (SCZ) GWAS and preserved those that could be mapped to both. Linkage disequilibrium (LD) clumping was conducted based on r^2^ > 0.0001 within a 1,000 kb window to select independent variants. To recruit enough instrumental variables (at least five), we used a lenient threshold for single nucleotide polymorphism (SNP) selection and identified eight SNPs for smoking initiation with *a P*-value of 5 × 10^−7^and five SNPs for onset age with *a P*-value of 1 × 10^−6^.

#### Genetic instruments for bidirectional MR

A bidirectional MR was conducted to estimate the causal effect of SCZ on smoking behaviors and examine whether an MR study supports the argument of self-medication in patients with SCZ. The data for bidirectional MR were taken from GWAS summary data of the Asian population with 18 independent SNPs of genome-wide significance (*P* < 5 × 10^−8^) for both smoking initiation and onset age.

#### MR analysis

In this study, five MR methods were applied as follows: inverse variance-weighted (IVW) (Burgess et al., 2013; Lawlor et al., 2008), weighted median (Bowden et al., 2016), weighted mode (Hartwig et al., 2017), Mendelian Randomization Pleiotropy Residual Sum and Outlier (MR-PRESSO) (Verbanck et al., 2018), and MR Egger (Bowden et al., 2015). IVW provides the best unbiased estimation in the absence of pleiotropy and under valid SNPs; hence, the main results for MR analysis are based on IVW method and the other four MR methods for the sensitivity analyses. We also examined pleiotropy using Egger intercept and heterogeneity using Cochran’s Q and Rücker’s Q.

### 2.4. Genetic correlation

Genetic correlation between two smoking traits and SCZ among East Asians was conducted to detect whether common genetic variants existed between these two phenotypes. The genetic correlation was measured using LD score regression (Bulik-Sullivan et al., 2015a, 2015b).

## 3. Results

### 3.1. Smoking GWAS in the Asian population

The Manhattan plot for two smoking traits for TWBB is shown in Figure 1. One novel locus was identified in TWBB (rs553874586) (Table 1). Rs553874586 in the *MMD* gene on chromosome 17 was associated with onset age (beta = −3.77, SE = 0.654, *P* = 8.08 × 10^−9^). In the GWAS meta-analysis of TWBB and BBJ, three SNPs met genome-wide significance, and all of them were novel (Table 1). Among these novel loci, variants rs79574881 at 8p22 (beta = −0.02, SE = 0.004, *P* = 2.84 × 10^−8^) and rs2156008 at 18q21.2 (beta = 0.01, SE = 0.002, *P* = 3.21 × 10^−8^) are associated with smoking initiation, and rs553874586 at 17q22 (beta = −3.77, SE = 0.654, *P* = 8.07 × 10^−9^) is associated with onset age.

**Figure 1.**
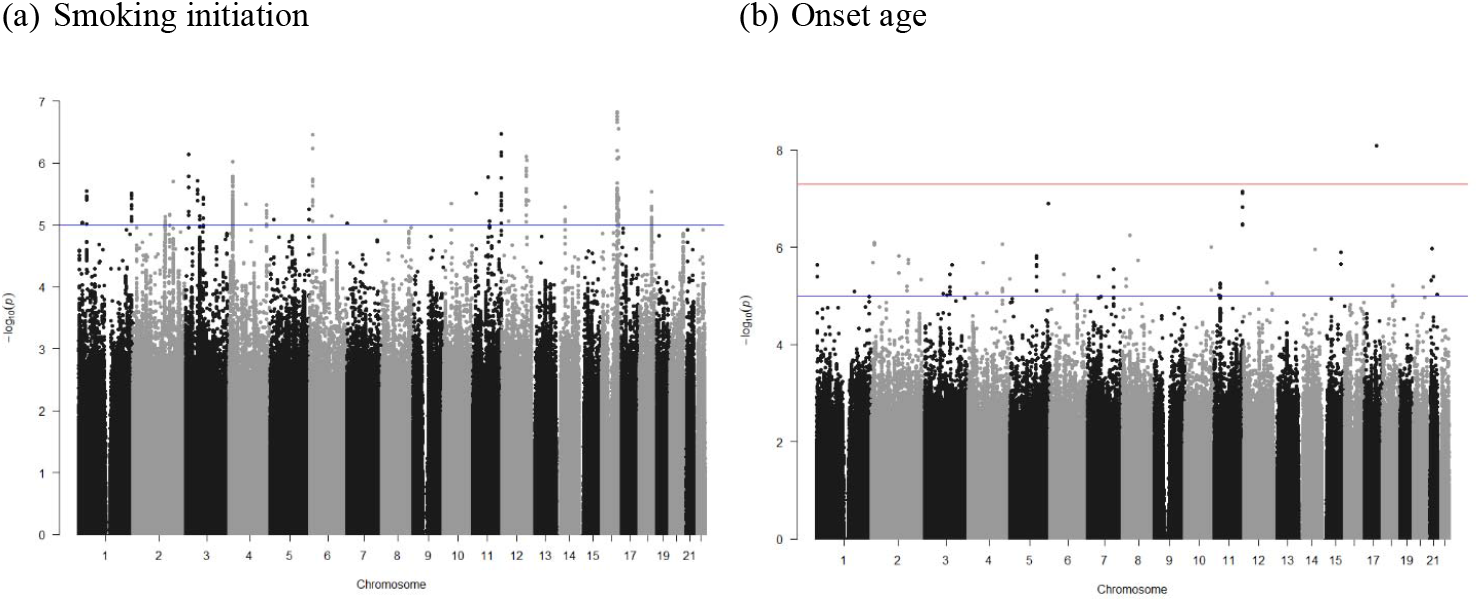
Manhattan plot for smoking behaviors among Taiwan Biobank data, including (a) smoking initiation, (b) onset age. The red line indicates the genome-wide significant level (*p*<5E-08) and the blue line indicates suggest significant level (*p*<1E-05).

### 3.2. Causal relationships of smoking and SCZ by MR study

Scatter plots of the causal estimation for smoking behaviors on SCZ is presented in Supplementary Figure 1, and forest plots are presented in Supplementary Figure 2. No heterogeneity was detected in the MR analysis for causality of the two smoking traits on SCZ (Supplementary Table 3).

The IVW method showed that smoking initiation was causally associated with an increased risk of SCZ with marginal significance (odds ratio [OR] = 4.00, 95% confidence interval [CI] = 0.89–18.01, *P* = 0.071) (Table 2). MR-PRESSO detected no outliers and no overall pleiotropy (*P*-value for MR Egger intercept = 0.99). Other MR methods provided similar estimates but did not reach statistical significance.

For causality of onset age of smoking on SCZ, the IVW method showed a marginal significance. A later onset age for smoking was causally associated with a lower risk of SCZ (OR for a per-year increase in onset = 0.96, 95% CI = 0.91–1.01, *P* = 0.098). There was no overall pleiotropy (*P*-value for MR Egger intercept = 0.68) (Table 2).

### 3.3. Bidirectional MR

We used Asia PGC data to identify genetic instruments for SCZ. The MR analyses did not support the causality of SCZ on the two smoking traits (see Supplementary Table 4).

### 3.4. Genetic correlation of smoking and SCZ

No significant genetic correlation was found between smoking behavior and SCZ among the Asian population. The genetic correlations were low, both for smoking initiation (r_g_ = −0.003, *P*= 0.94) and onset age (r_g_ = 0.102, *P* = 0.17) and SCZ.

## 4. Discussion

Using improved-power GWAS data from the EAS population, we detected three novel variants related to smoking initiation and onset age. The MR analysis provided evidence for the harmful effect of smoking initiation and earlier onset age on SCZ in the EAS population, which was in line with previous evidence in the European population.

Three novel variants were detected in relation to smoking behaviors in a meta-analysis of the Asian population. Among these SNPs, rs79574881 and rs2156008 were associated with smoking initiation. Rs79574881 is located on *the DLC1* gene, which encodes a GTPase-activating protein. It is a tumor suppressor gene and has been reported to be related to prostate, lung, and breast cancers (Tripathi et al., 2014; Wang et al., 2020). Further, variants in *the DLC1* gene have also been associated with nicotine dependence (Gelernter et al., 2015). Rs2156008 is located on *the TCF4* gene, which is a protein-coding gene related to nervous system development. Many SNPs in *the TCF4* gene are associated with SCZ or SCZ endophenotypes (Quednow et al., 2014). Moreover, the gene effects were modulated by smoking behavior, where heavy-smokers showed stronger gene effects on the SCZ endophenotype than light-smokers and non-smokers (Quednow et al., 2012).

Rs553874586 is a novel SNP associated with the onset age of smoking in our study. This SNP is located on *the MMD* gene, which is a protein-coding gene, and its molecular functions include protein kinase activity and signaling receptor activity.

Consistent and strong evidence has been reported for the causal effect of smoking initiation on SCZ among European-origin individuals (Barkhuizen et al., 2021; Gage et al., 2017; Wootton et al., 2020; Yuan et al., 2020); however, this issue has been less discussed in the Asian populations. Only one MR analysis in Asian participants with limited statistical power reported no significant causal association between smoking and SCZ (Chen et al., 2021). Concerning that the biological mechanism should be presented in different populations if the causal effect between two phenotypes is truly existence, we increased the sample size for exposure and outcome variables and found that smoking initiation showed a marginal causal association with SCZ. Hence, partial evidence was found for the causal effect of smoking initiation on SCZ among Asian populations for a sufficient sample size, which corroborates with previous findings in the European population. In addition to previous MR studies, our results are supported by studies of various study designs. One retrospective study reported that 90% of patients with SCZ started smoking before the onset of their disease (Kelly & McCreadie, 1999), and longitudinal studies have pointed out that cigarette smoking increases the risk of developing SCZ and consistently reported a dose-response relationship of smoking quantity (Kendler et al., 2015; McGrath et al., 2016; Mustonen et al., 2018; Weiser et al., 2004). A previous MR study with 632,802 Caucasian population of smoking initiation GWAS also supported our finding, which indicated a causal role of smoking initiation in many psychiatric disorders, including SCZ, depression, and bipolar disorder (Barkhuizen et al., 2021).

Previous studies have provided evidence of shared genetic variants between smoking and SCZ. For example, researchers found that the risk of SCZ increased with an increase in the polygenic risk score of plasma cotinine concentration, and a higher genetic score for SCZ increased the risk of smoking behaviors (Chen et al., 2016). Another study also reported a positive correlation between polygenic risk scores between regular smoking and SCZ (Harper et al., 2021). This implies some shared genetic liability between SCZ and smoking behaviors, which also describes the possible biological mechanism between smoking behaviors and SCZ. However, the genetic correlation between these two phenotypes did not achieve statistical significance in our study, which may have resulted from LD varying across different populations (Hill, 2013).

Brain structure may explain the causality of smoking and SCZ. For example, researchers have observed that event-related electroencephalogram activity (P3/δ component) showed a negative correlation with SCZ polygenic risk scores and a prefrontal control component (θ/antisaccade) was negatively correlated with number of drinks per week and regular smoking polygenic risk scores (Harper et al., 2021). Because the brain structure and function in different brain regions are closely related, this finding provides evidence that brain structure may simultaneously affect the SCZ status and smoking behaviors.

Using a genetic approach to investigate the causality between smoking behaviors and SCZ can avoid possible confounders and ensure the temporal sequence. Although investigating an Asian population provided an opportunity to investigate causality particularly in this population, the sample size for smoking GWAS was insufficient to provide enough instruments that met genome-wide significance (*P* = 5 × 10^−8^). Instead, we used a wider P-value threshold at *P* = 5 × 10^−7^ for smoking initiation and *P* = 1 × 10^−6^ for onset age to include sufficient SNPs. This may recruit weak instrumental variables and lead to bias in MR analysis. Furthermore, we only included two smoking behaviors (smoking initiation and onset age of smoking) in the analysis; hence, the causal effect of a wide range of smoking traits on SCZ needs to be examined further.

In conclusion, this study identified two novel SNPs related to smoking initiation and one novel SNP related to the onset age. We also provided evidence for the marginal causal effect of smoking initiation on SCZ, which corresponds to the longitudinal and cross-sectional studies of the association between smoking and SCZ (de Leon, 1996; Kelly & McCreadie, 1999; Weiser et al., 2004). SCZ is a complex trait, and MR analysis can help identify possible causal risk factors. Future efforts to elucidate the mechanisms underlying the association between smoking and SCZ are needed and may help early prevention.

## Data Availability

All data produced in the present study are available upon reasonable request to the corresponding author

## Acknowledgement

This work was supported by the Taiwanese National Health Research Institutes (NHRI-EX109-10931PI, NHRI-EX110-10931PI, and NHRI-EX111-10931PI) and China Medical University (CMU110-MF-79).

## Data availability

GWAS summary results for schizophrenia are available on the PGC website https://www.med.unc.edu/pgc/. GWAS summary results for smoking behaviors are available on the website of Biobank Japan http://jenger.riken.jp/en/result.

## Reference

Barkhuizen, W., Dudbridge, F., & Ronald, A. (2021). Genetic overlap and causal associations between smoking behaviours and mental health. Sci Rep, 11(1), 14871.

Bowden, J., Davey Smith, G., & Burgess, S. (2015). Mendelian randomization with invalid instruments: effect estimation and bias detection through Egger regression. Int J Epidemiol, 44(2), 512–525.

Bowden, J., Davey Smith, G., Haycock, P. C., & Burgess, S. (2016). Consistent Estimation in Mendelian Randomization with Some Invalid Instruments Using a Weighted Median Estimator. Genet Epidemiol, 40(4), 304–314.

Bulik-Sullivan, Brendan, Finucane Hilary K, Anttila, Verneri, Gusev, Alexander, Day Felix R, Loh, Po-Ru, et al. (2015a). An atlas of genetic correlations across human diseases and traits. Nat Genet, 47(11), 1236–1241.

Bulik-Sullivan, Brendan K, Loh, Po-Ru, Finucane Hilary K, Ripke, Stephan, Yang, Jian, Patterson, Nick, et al. (2015b). LD Score regression distinguishes confounding from polygenicity in genome-wide association studies. Nat Genet, 47(3), 291–295.

Burgess, S., Butterworth, A., & Thompson, SG. (2013). Mendelian randomization analysis with multiple genetic variants using summarized data. Genet Epidemiol, 37(7), 658–665.

Chen, J., Bacanu, SA., Yu, H., Zhao, Z., Jia, P., Kendler, KS., et al. (2016). Genetic relationship between schizophrenia and nicotine dependence. Sci Rep. doi:10.1038/srep25671.

Chen, J., Chen, R., Xiang, S., Li, N., Gao, C., Wu, C., et al. (2021). Cigarette smoking and schizophrenia: Mendelian randomisation study. Br J Psychiatry, 218(2), 98–103.

de Leon, J. (1996). Smoking and vulnerability for schizophrenia. Schizophr Bull, 22(3), 405–409.

Gage, SH., Jones, HJ., Taylor, AE., Burgess, S., Zammit, S., & Munafò, MR. (2017). Investigating causality in associations between smoking initiation and schizophrenia using Mendelian randomization. Sci Rep, 7, 40653.

Gelernter, J., Kranzler, HR., Sherva, R., Almasy, L., Herman, AI., Koesterer, R., et al. (2015). Genome-Wide Association Study of Nicotine Dependence in American Populations: Identification of Novel Risk Loci in Both African-Americans and European-Americans. Biol Psychiatry, 77(5), 493–503.

Gurillo, P., Jauhar, S., Murray, RM., & MacCabe, JH. (2015). Does tobacco use cause psychosis? Systematic review and meta-analysis. Lancet Psychiatry, 2(8), 718–725.

Harper, J., Liu, M., Malone, SM., McGue, M., Iacono, WG., & Vrieze, SI. (2021). Using multivariate endophenotypes to identify psychophysiological mechanisms associated with polygenic scores for substance use, schizophrenia, and education attainment. Psychol Med, 1–11. doi:10.1017/S0033291721000763

Hartwig, FP., Davey Smith, G., & Bowden, J. (2017). Robust inference in summary data Mendelian randomization via the zero modal pleiotropy assumption. Int J Epidemiol, 46(6), 1985–1998.

Hill, WG. (2013). Brenner’s Encyclopedia of Genetics (S. Maloy & K. Hughes Eds. 2nd ed.): Elsevier.

Hunter, A., Murray, R., Asher, L., & Leonardi-Bee, J. (2020). The Effects of Tobacco Smoking, and Prenatal Tobacco Smoke Exposure, on Risk of Schizophrenia: A Systematic Review and Meta-Analysis. Nicotine Tob Res, 22(1), 3–10.

Kelly, C., & McCreadie, RG. (1999). Smoking habits, current symptoms, and premorbid characteristics of schizophrenic patients in Nithsdale, Scotland. Am J Psychiatry, 156(11), 1751–1757.

Kendler, KS., Lonn, SL., Sundquist, J., & Sundquist, K. (2015). Smoking and schizophrenia in population cohorts of Swedish women and men: a prospective co-relative control study. Am J Psychiatry, 172(11), 1092–1100.

Lam, M., Chen, C. Y., Li, Z., Martin, A. R., Bryois, J., Ma, X., et al. (2019). Comparative genetic architectures of schizophrenia in East Asian and European populations. Nat Genet, 51(12), 1670–1678. doi:10.1038/s41588-019-0512-x

Lawlor, DA., Harbord, RM., Sterne, JA., Timpson, N., & Davey Smith, G. (2008). Mendelian randomization: using genes as instruments for making causal inferences in epidemiology. Stat Med, 27(8), 1133–1163.

Matoba, N., Akiyama, M., Ishigaki, K., Kanai, M., Takahashi, A., Momozawa, Y., et al. (2019). GWAS of smoking behaviour in 165,436 Japanese people reveals seven new loci and shared genetic architecture. Nat Hum Behav, 3(5), 471–477.

McGrath, JJ., Alati, R., Clavarino, A., Williams, GM., Bor, W., Najman, JM., et al. (2016). Age at first tobacco use and risk of subsequent psychosis-related outcomes: A birth cohort study. Aust N Z J Psychiatry, 50(6), 577–583.

Moudi, A., Dashtgard, A., Salehiniya, H., Sadat Katebi, M., Reza Razmara, M., & Reza Jani, M. (2018). The relationship between health-promoting lifestyle and sleep quality in postmenopausal women. Biomedicine (Taipei), 8(2), 11.

Mustonen, A., Ahokas, T., Nordström, T., Murray, GK., Mäki, P., Jääskeläinen, E., et al. (2018). Smokin’ hot: adolescent smoking and the risk of psychosis. Acta Psychiatr Scand, 138(1), 5–14.

Quednow, BB., Brinkmeyer, J., Mobascher, A., Nothnagel, M., Musso, F., Gründer, G., et al. (2012). Schizophrenia risk polymorphisms in the TCF4 gene interact with smoking in the modulation of auditory sensory gating. Proc Natl Acad Sci U S A, 109(16), 6271–6276.

Quednow, BB., Brzózka, MM., & Rossner, MJ. (2014). Transcription factor 4 (TCF4) and schizophrenia: integrating the animal and the human perspective. Cell Mol Life Sci, 71(15), 2815–2835.

Teramoto, M., Iso, H., Wakai, K., & Tamakoshi, A. (2021). Secondhand Smoke Exposure during Childhood and Cancer Mortality in Adulthood among Never Smokers: the Japan Collaborative Cohort Study for Evaluation of Cancer Risk. Am J Epidemiol(kwab284). doi:10.1093/aje/kwab284

Tripathi, V., Popescu, NC., & Zimonjic, DB. (2014). DLC1 induces expression of E-cadherin in prostate cancer cells through Rho pathway and suppresses invasion. Oncogene, 33(6), 724–733.

Verbanck, M, Chen, CY, Neale, B, & Do, R. (2018). Detection of widespread horizontal pleiotropy in causal relationships inferred from Mendelian randomization between complex traits and diseases. Nat Genet, 50(5), 693–698.

Wang, D., Qian, X., Sanchez-Solana, B., Tripathi, BK., Durkin, ME., & Lowy, DR. (2020). Cancer-Associated Point Mutations in the DLC1 Tumor Suppressor and Other Rho-GAPs Occur Frequently and Are Associated with Decreased Function. Cancer Res, 80(17), 3568–3579.

Wang, SH., Lai, RY., Lee, YC., Su, MH., Chen, CY., Hsiao, PC., et al. (2019). Association between polygenic liability for schizophrenia and substance involvement: A nationwide population-based study in Taiwan. Genes Brain Behav, 19(5), e12639.

Weiser, Mark, Reichenberg, Abraham, Grotto, Itamar, Yasvitzky, Ross, Rabinowitz, Jonathan, Lubin, Gad, et al. (2004). Higher rates of cigarette smoking in male adolescents before the onset of schizophrenia: a historical-prospective cohort study. Am J Psychiatry, 161, 1219–1223.

Whiteford, Harvey A, Degenhardt, Louisa, Rehm Jürgen, Baxter, Amanda J, Ferrari Alize J, Erskine Holly E, et al. (2013). Global burden of disease attributable to mental and substance use disorders: findings from the Global Burden of Disease Study 2010. Lancet, 382(9904), 1575–1586.

Winterer, G. (2010). Why do patients with schizophrenia smoke? Curr Opin Pharmacol, 23(2), 112–119.

Wootton, RE., Richmond, RC., Stuijfzand, BG., Lawn, RB., Sallis, HM., Taylor, GMJ., et al. (2020). Evidence for causal effects of lifetime smoking on risk for depression and schizophrenia: a Mendelian randomisation study. Psychol Med, 50(14), 2435–2443.

Yuan, S., Yao, H., & Larsson, SC. (2020). Associations of cigarette smoking with psychiatric disorders: evidence from a two-sample Mendelian randomization study. Sci Rep, 10(1), 13807.

